# Impact of neoadjuvant chemoradiotherapy on post-operative clinically significant pancreatic fistula – A systemic review and updated meta-analysis

**DOI:** 10.1101/2020.12.14.20248164

**Authors:** Bhavin Vasavada, Hardik Patel

## Abstract

**Background:** Pancreatic fistula is one of the main complications after pancreatic surgery and the leading cause of morbidity and mortality after pancreatic surgery. There are many pieces of evidence emerging out from retrospective studies and metanalysis that neoadjuvant chemoradiation decreases rates of clinically significant postoperative pancreatic fistula.

**Aims and objectives:** The primary aim of our analysis was to do a systemic review and updated meta-analysis of literature published in the last 10 years and look for the association of neoadjuvant chemoradiation and risk of subsequent clinically significant pancreatic fistula.

**Methods:** EMBASE, MEDLINE, and the Cochrane Database were searched for Studies comparing outcomes in patients receiving neoadjuvant chemoradiotherapy first with those patients who received surgery first in case of pancreatic cancer. A systemic review and Metanalysis were done according to MOOSE and PRISMA guidelines. Heterogeneity was measured using Q tests and I2, and p < 0.10 was determined as significant. The Odds ratios (OR) and Risk Ratios (RR) were calculated for dichotomous data as per the requirement, and weighted mean differences (WMD) were used for continuous variables. Nonrandomized trials were accessed for bias using the New Castle Ottawa scale. Publication bias was studied using funnel plots. The meta-analysis was conducted using Open Review Manager 5.4.

**Results:** Twenty-six studies including 17021 patients finally included in the analysis. 339 patients out of a total of 3386 developed clinically significant pancreatic fistula in the neoadjuvant first group. 2342 patients out of 13335 patients developed clinically significant pancreatic fistula in the surgery first group. Neoadjuvant treatment significantly reduced the risk of subsequent clinically significant pancreatic fistula. (p= <0.0001). The number of patients with soft pancreas was significantly higher in the surgery first group. (p <0.0001). Pancreatic duct diameter mentioned in only two studies but there was no significant difference between both groups. [p=1].Blood loss was significantly more in the surgery first group. [p <0.0001]. There was no difference in pancreaticoduodenectomy or distal pancreatectomy performed between both groups. (p=0.82). There was no difference in the number of borderline resectable pancreatic tumors between both groups. (p= 0.34). There was no difference in overall grade 3/grade 4 complications rate between both groups. (p= 0.39).

**Conclusion:** Neoadjuvant treatments may be responsible for the lower rates of clinically significant pancreatic fistula after subsequent surgery.

## Background

Pancreatic fistula is one of the main complications after pancreatic surgery and the leading cause of morbidity and mortality after pancreatic surgery. In spite of all the scientific progress pancreatic surgery is still associated with very high complication rates. [1,2,3,4].Pancreaticoduodenectomy is generally performed for pancreatic ductal adenocarcinoma in the head region and distal pancreatectomy for the body and tail region. R0 resection is necessary to increase the chances of long-term survival. When a tumor invades the superior mesenteric vein or superior mesenteric artery it is sometimes difficult to achieve R0 resection, which can hamper chances of long-term survival. These tumors are defined as borderline resectable tumors. [5,6,7] Consensus definition of borderline resectable pancreatic cancer includes anatomical considerations(contact with <180 degrees of the superior mesenteric artery and/or celiac artery, short segment contact with the common hepatic artery, and contact or occlusion with the superior mesenteric vein□portal vein confluence with adequate vein proximal and distal for reconstruction, high□risk biologic features, and patient performance status. [8,9]. Some studies have shown that neoadjuvant chemoradiation improves R0 resection rates in borderline resectable pancreatic tumors. the current National Comprehensive Cancer Network guidelines recommend neoadjuvant treatment for borderline resectable pancreatic tumors. [10,11]

Now there are many shreds of evidence emerging out from retrospective studies and metanalysis that neoadjuvant chemoradiation decreases rates of clinically significant postoperative pancreatic fistula. [13,14,15]

## AIMS AND OBJECTIVES

The primary aim of our analysis was to do a systemic review and updated meta-analysis of literature published in the last 10 years and look for the association of neoadjuvant chemoradiation and risk of subsequent clinically significant pancreatic fistula. We also looked for other components of fistula risk score between the neoadjuvant chemoradiotherapy group and the surgery first group.[16]

## Material and Methods

EMBASE, MEDLINE, and the Cochrane Database were searched for Studies comparing outcomes in patients receiving neoadjuvant chemoradiotherapy first with those patients who received surgery first in case of pancreatic cancer. We used keywords like “neoadjuvant chemoradiotherapy” “pancreas cancer” “Surgery first” “borderline resectable pancreas cancer”. Only English language articles selected. Two independent authors extracted the data (B.V and H.P). Discussions and mutual understanding resolved any disagreements. A systemic review and Metanalysis were done according to MOOSE and PRISMA guidelines. [17,18]

### Statistical analysis

The meta-analysis was conducted using Open Review Manager 5.4. Heterogeneity was measured using Q tests and I2, and p < 0.10 was determined as significant [19]. If there was no or low heterogeneity (I2 < 25%), then the fixed-effects model was used. Otherwise, the random-effects model was used. The Odds ratios (OR) and Risk Ratios (RR) were calculated for dichotomous data as per the requirement, and weighted mean differences (WMD) were used for continuous variables. Both differences were presented with 95% CI. For continuous variables, if data were presented with medians and ranges, then we calculated the means and Standard deviations according to Hozo et al. [20]. If the study presented the median and inter-quartile range, the median was treated as the mean, and the interquartile ranges were calculated using 1.35 SDs, as described in the Cochrane handbook.

### Assessment of Bias

Nonrandomized trials were accessed using the New Castle Ottawa scale [21] and randomized trials were accessed using the Cochrane handbook. [22]. Publication bias was studied using funnel plots.[Supplement Figure 1]. all relevant data were available from the original publications. We did not impute any missing outcome data. We presented the overall quality of the evidence for each outcome according to the GRADE approach, which takes into account issues not only related to internal validity (risk of bias, inconsistency, imprecision, publication bias) but also to external validity such as directness of results. [24].We created a ‘Summary of findings’ table based on the methods described in the Cochrane Handbook for Systematic Reviews of Intervention. [25]

### Definitions

We defined clinically significant pancreatic fistula as per the ISGPS definition. [24] and we included any neoadjuvant therapy in the neoadjuvant therapy group.

### Inclusion criteria of studies

* Full-text articles
* Articles containing both neoadjuvant chemoradiotherapy group and surgery first group
* Studied published in the last 10 years.
* English language articles
* Studies which included pancreatic fistula analysis
* Study involving pancreas ductal adenocarcinoma only.

### Exclusion criteria

* Articles where full texts could not be obtained
* Duplicate studies
* Studies that used duplicate data
* Articles without the English language

## Results

### Study Selection

The study selection process is described in Figure: 1 as per PRISMA guidelines. Twenty-six studies including 17021 patients finally included in the analysis. [26-51]. Study characteristics are described in Table 1. 3386 patients were in the neoadjuvant chemoradiation group and 13335 patients were in the surgery first group. The risk of bias summary is described in Figure 2.

**TABLE 1.** STUDY CHARACTERISTICS

| STYDY ID | YEAR | TIME PERIOD | TYPE OF STUDY | TOTAL PATIENT IN NEOADJUVANT ARM | TOTAL PATIENT IN SURGERY FIRST ARM |
| --- | --- | --- | --- | --- | --- |
| Araujo et al. | 2013 | 2001-2011 | Retrospective cohort | 29 | 29 |
| Barenboim et al. | 2008 | 2014-2017 | Retrospective cohort | 23 | 47 |
| Cloyd et al. | 2017 | 1999-2014 | Retrospective |  | 99 |
|  |  |  | cohort | 43 |  |
| Cooper et al. | 2015 | 2011-2012 | Retrospective cohort | 199 | 1363 |
| Denost et al. | 2013 | 2004-2009 | Retrospective cohort | 39 | 72 |
| Ellis et al. | 2019 | 2014-2017 | Retrospective cohort | 1864 | 8158 |
| Epemboym et al. | 2013 | 1992-2011 | Retrospective cohort | 143 | 363 |
| Jason et al | 2016 | 2011-2015 | Prospective cohort | 137 | 72 |
| Jiang et al. | 2013 | 2004-2010 | Retrospective cohort | 112 | 120 |
| Lee et al. | 2015 | 2000-2013 | Retrospective cohort | 30 | 28 |
| Lof et al. | 2019 | 2007-2015 | Retrospective cohort | 136 | 1100 |
| Marchegiani et al. | 2017 | 2014-2016 | Retrospective cohort | 99 | 206 |
| Mellon et al. | 2016 | 2000-2012 | Retrospective cohort | 61 | 241 |
| Nadya et al. | 2020 | 2011-2017 | Prospective cohort | 76 | 58 |
| Nicolas et al. | 2020 | 2012-2017 | Retrospective | 164 | 284 |
|  |  |  | cohort |  |  |
| Paye et al. | 2014 | 2004-2009 | Retrospective cohort | 39 | 248 |
| Pecorelli et al. | 2012 | 2003-2011 | Retrospective cohort | 50 | 100 |
| Pecorelli et al. | 2019 | 2015-2018 | Retrospective cohort | 95 | 190 |
| PREOPANC TRIAL | 2020 | 2013-2017 | Randomised control trial | 66 | 98 |
| SHO et al. | 2012 | 2008-2011 | Retrospective cohort | 61 | 71 |
| TAKAHASHI et al. | 2011 | 2004-2007 | Retrospective cohort | 28 | 30 |
| Tashiro et al. | 2018 | 2010-2017 | Retrospective cohort | 67 | 94 |
| Timmermann et al | 2019 | 2016-2018 | Retrospective cohort | 33 | 99 |
| Uchida et al. | 2020 | 2012-2019 | Retrospective cohort | 52 | 148 |
| Yoshitomi et al. | 2018 | 2010-2016 | Retrospective cohort | 31 | 7 |
| Yoshiya et al. | 2019 | Not available | Retrospective cohort | 9 | 11 |

**Table 2:**
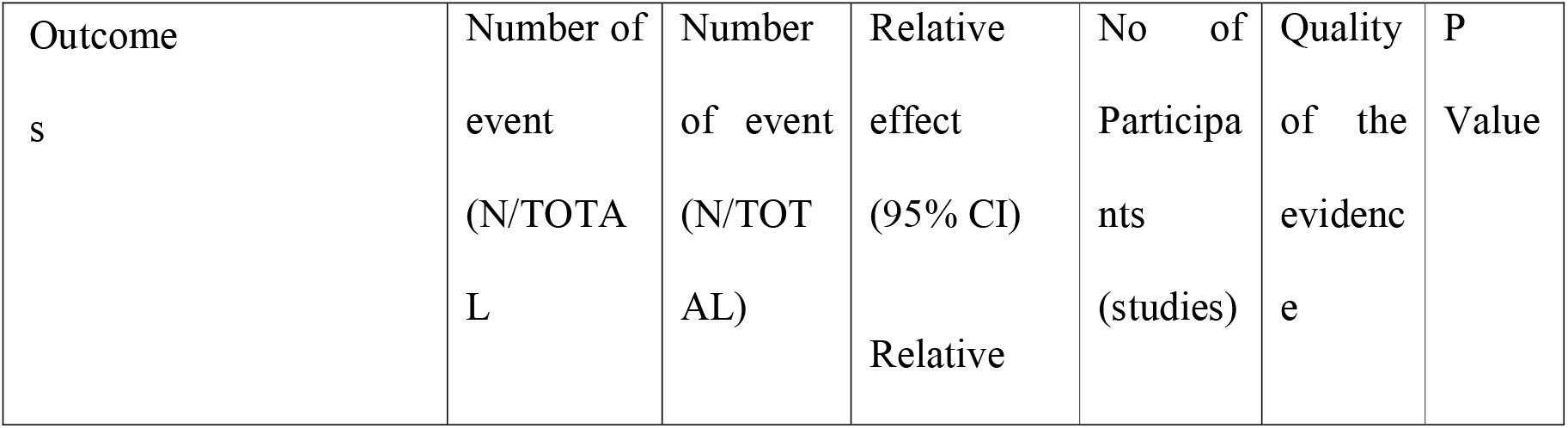
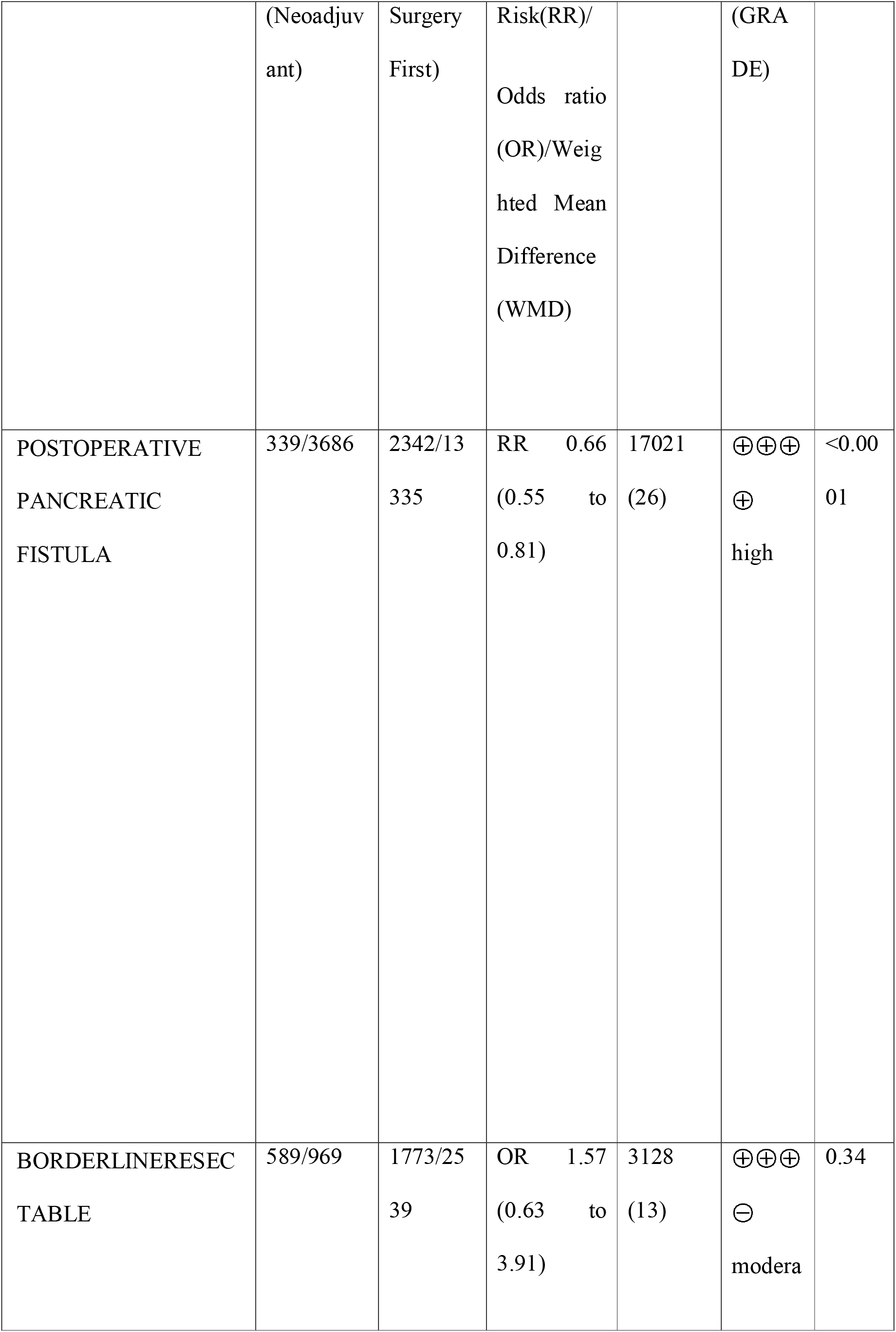
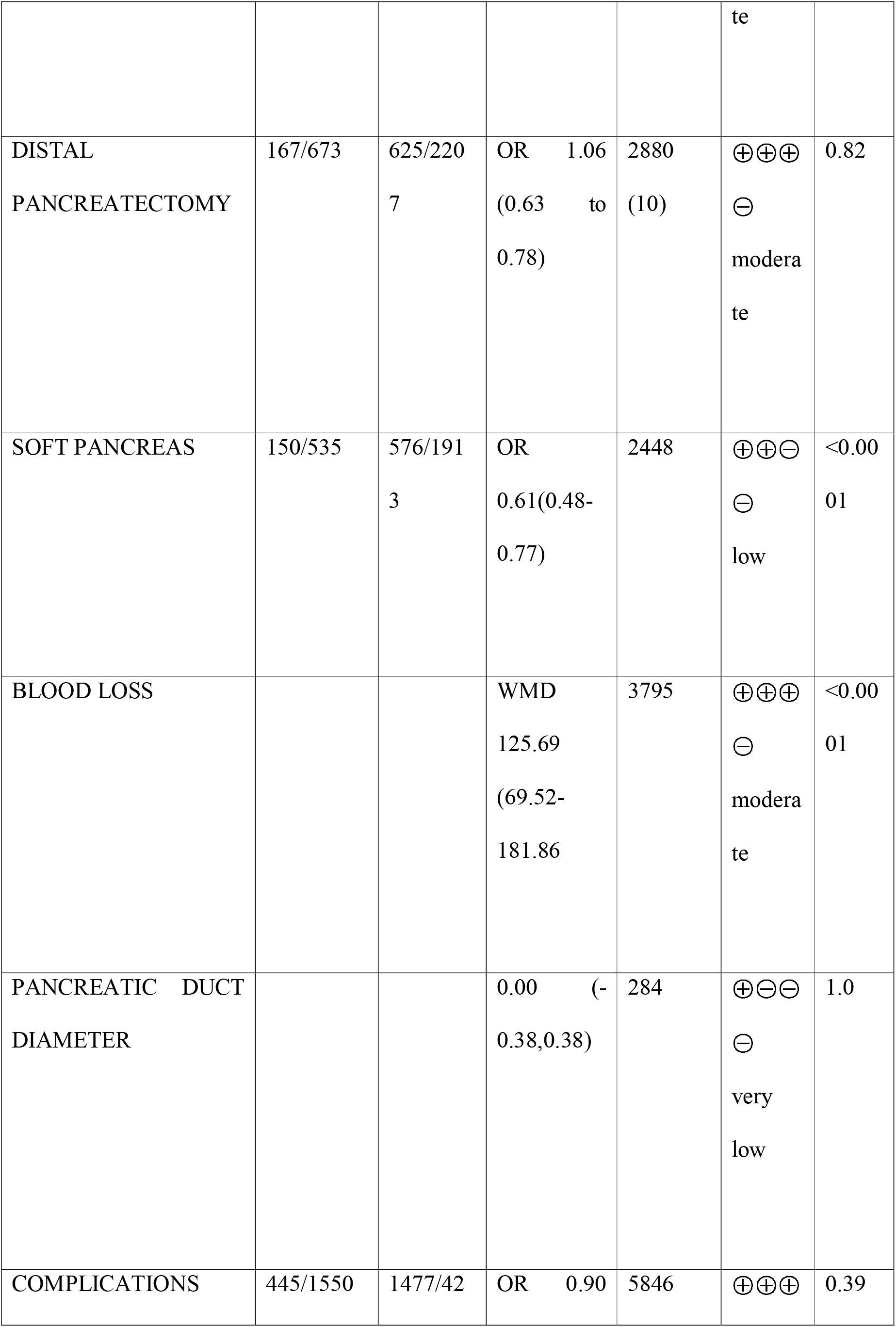
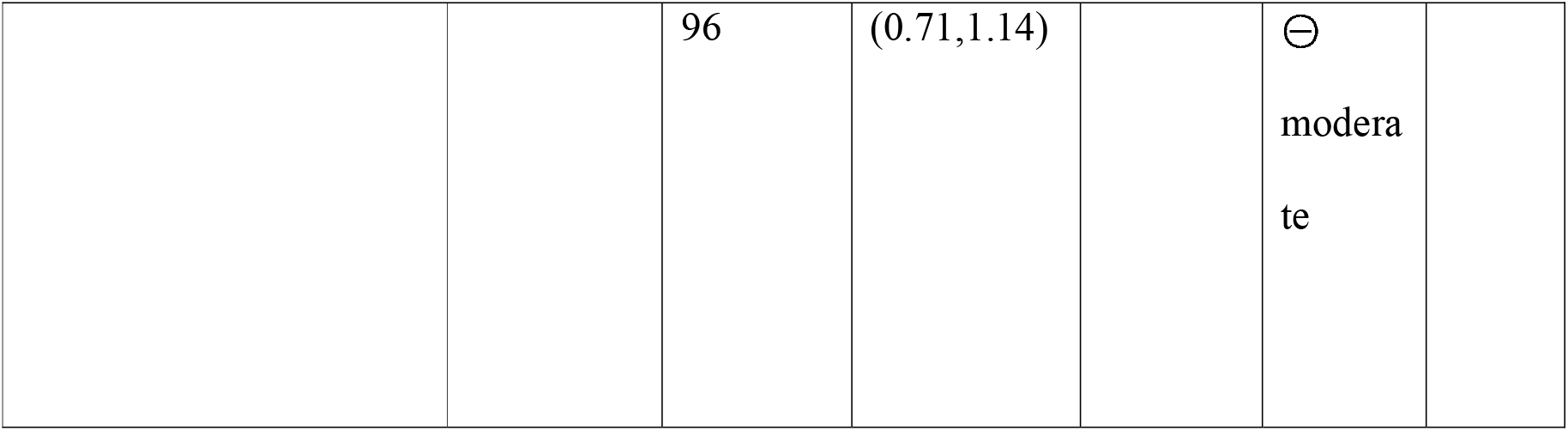
Summary of findings: Neoadjuvant Treatment compared with Surgery first for Pancreatic cancer Patient or population: Resectable/Borderline resectable with pancreatic Adenocarcinoma Intervention: Neoadjuvant Treatment Comparison: Surgery First. GRADE Working Group grades of evidence **High quality:** Further research is very unlikely to change our confidence in the estimate of effect. **Moderate quality:** Further research is likely to have an important impact on our confidence in the estimate of effect and may change the estimate. **Low quality:** Further research is very likely to have an important impact on our confidence in the estimate of effect and is likely to change the estimate. **Very low quality:** We are very uncertain about the estimate.

**Figure 1:**
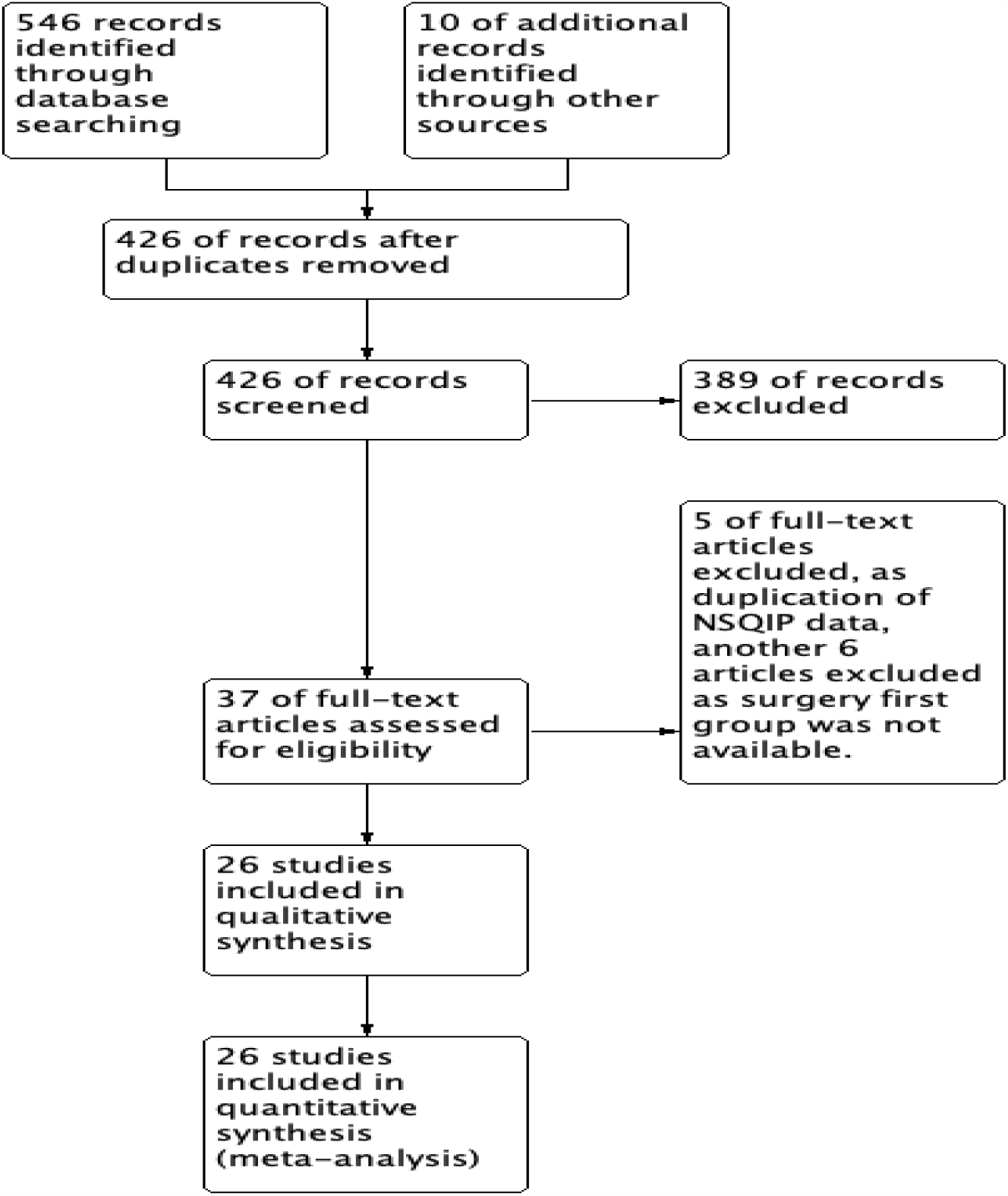
PRISMA flow chart.

**FIGURE 2.**
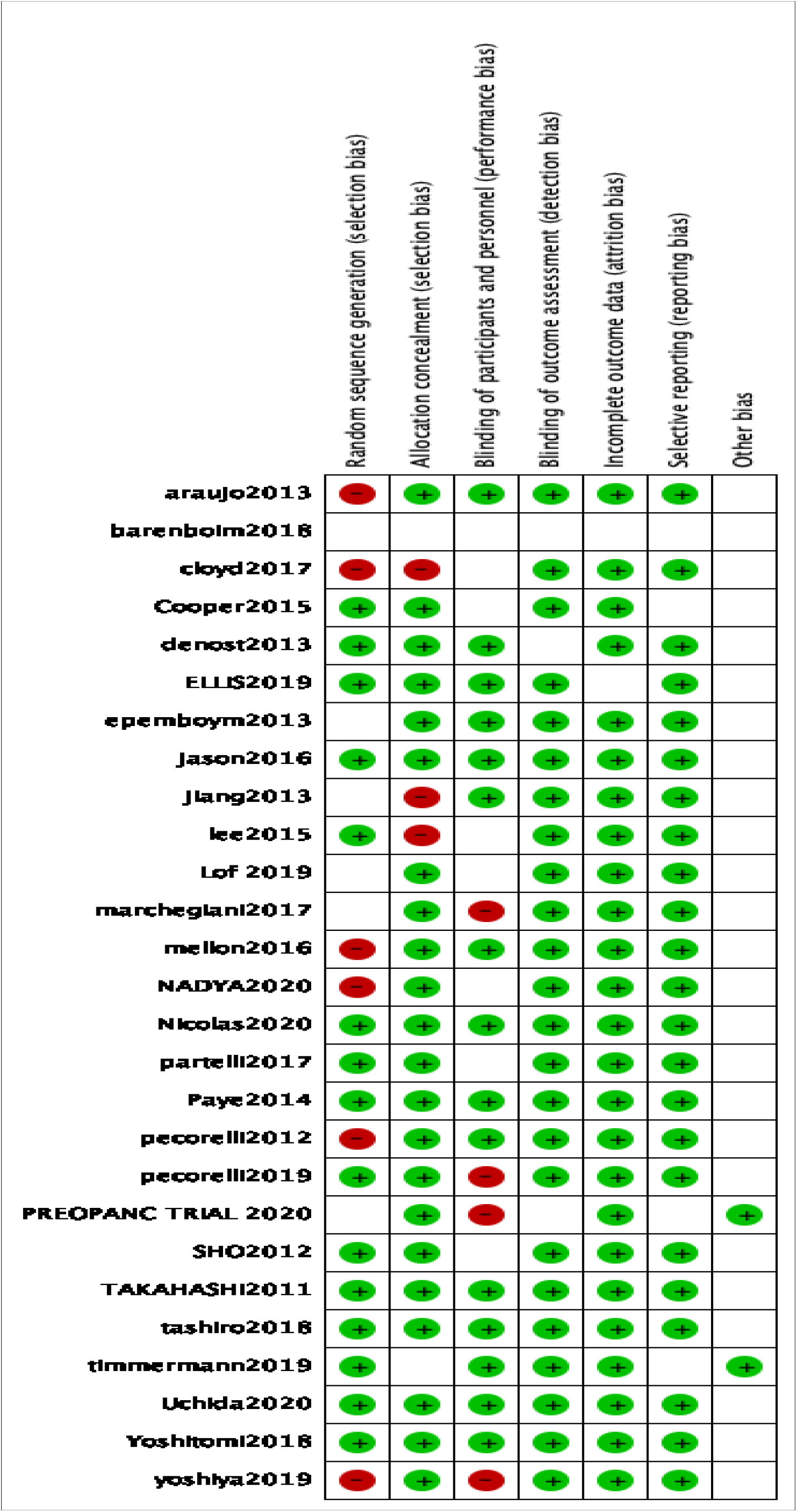
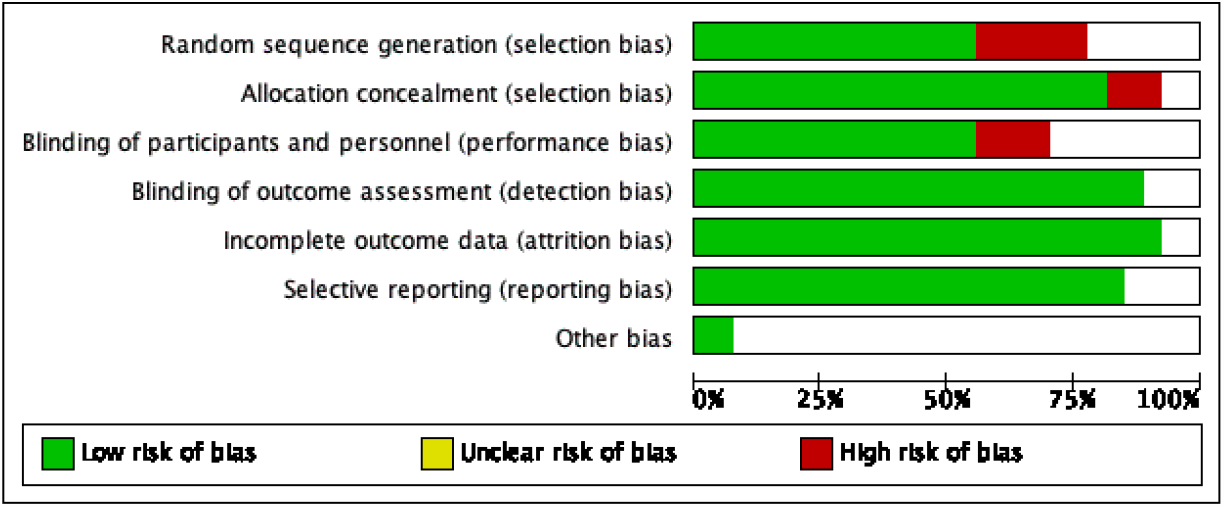
(a) Risk of bias summary: review authors’ judgements about each risk of bias item for each included study (b) Risk of bias graph: review authors’ judgements about each risk of bias item presented as percentages across all included studies.

### Comparisons of pancreatic fistulas between neoadjuvant first and surgery first

339 patients out of a total of 3386 developed clinically significant pancreatic fistula in the neoadjuvant first group. 2342 patients out of 13335 patients developed clinically significant pancreatic fistula in the surgery first group. Neoadjuvant treatment significantly reduced the risk of subsequent clinically significant pancreatic fistula. (p= <0.0001, Risk ratio 0.66, 95% confidence interval of risk ratio 0.55-0.81). [Figure 3]

**Figure 3.**
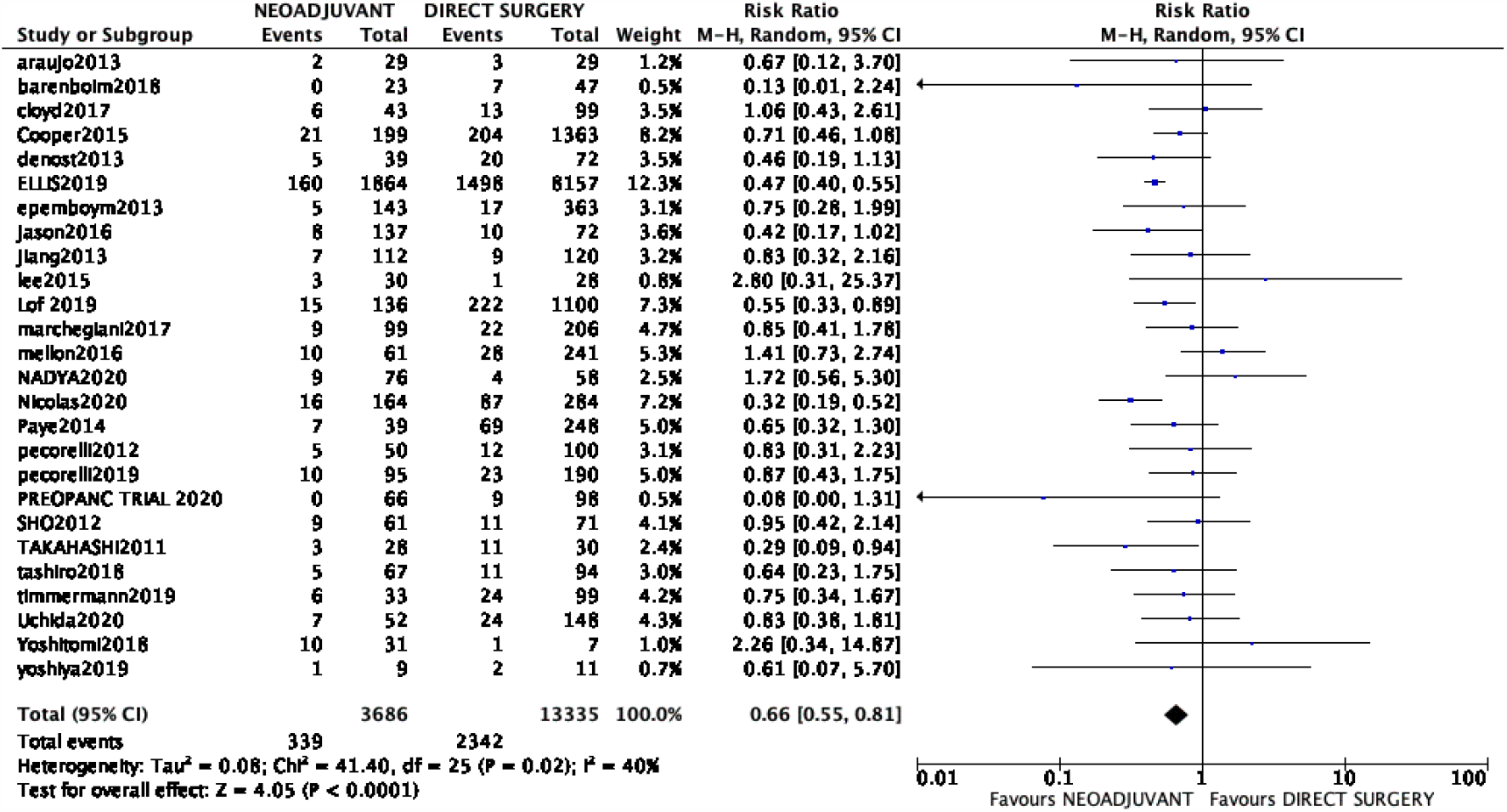
Forest plot of comparison: 1 Neoadjuvant chemoradiotherapy vs direct surgery in pancreatic cancer for postoperative pancreatic fistula.

### Components of Intraoperative fistula risk score

We also compared other components of fistula risk scores like gland textures (soft or not), blood loss, or duct diameter. The number of patients with soft pancreas was significantly higher in the surgery first group. (p <0.0001). Pancreatic duct diameter mentioned in only two studies but there was no significant difference between both groups. [p=1].Blood loss was significantly more in the surgery first group.[Mean difference 125.69 ml, 95% confidence interval 69.52-181.86 ml, p <0.0001] [Figure 4]. We included studies with pancreatic ductal adenocarcinomas only, so histology was similar in both the groups.

**Figure 4.**
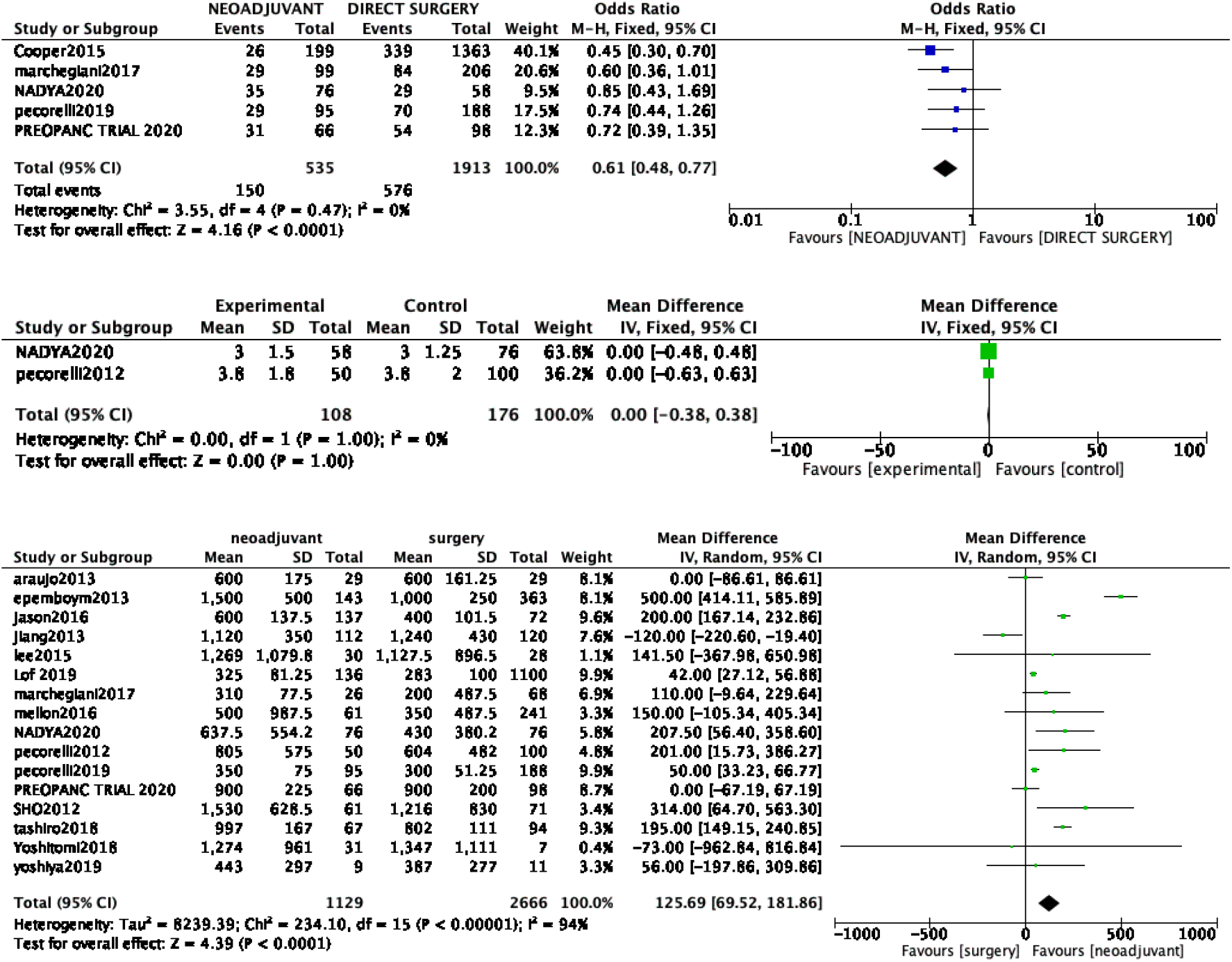
Forest plot of comparison (a) SOFT PANCREAS (b) pancreatic duct diameter. (c) Blood loss

### Type of surgeries, borderline resectable pancreatic tumor, and overall grade 3/grade 4 complications

There was no difference in pancreaticoduodenectomy or distal pancreatectomy performed between both groups. (p=0.82). There was no difference in the number of borderline resectable pancreatic tumors between both groups. (p= 0.34). There was no difference in overall grade 3/grade 4 complications rate between both groups. (p= 0.39). [Figure 5].

**Figure 5.**
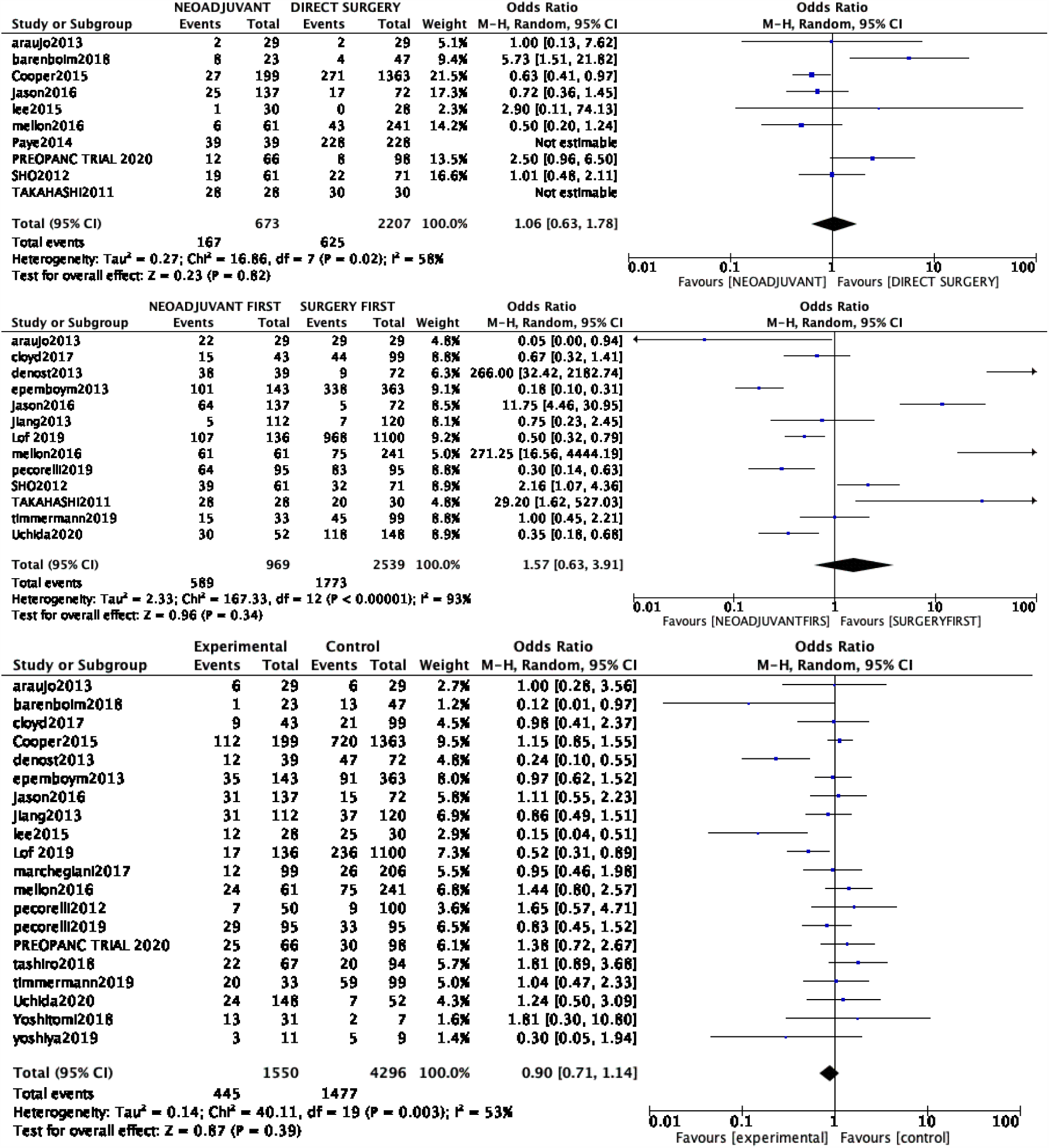
Forest plot of comparison (a) distal pancreatectomy (b) borderline resectable pancreatic tumours (c) overall complications.

## DISCUSSION

Neoadjuvant chemoradiotherapy is increasingly being used in pancreatic cancer to achieve increase R0 rates, NCCN guidelines also recommend neoadjuvant chemoradiotherapy as a part of the treatment protocol. [10,11]. Recently published meta-analysis has shown increased overall survival in such patients. [12] however, there was some concern about increased complication rates following neoadjuvant chemoradiotherapy protocol. The PREOPANC randomized control trial [49] showed that neoadjuvant protocol did not increase complication rates.

The PREOPANC trial and many other studies showed [49,50,51] that neoadjuvant chemoradiotherapy decreased postoperative pancreatic fistula rates also. These findings have been confirmed with systemic reviews and meta-analysis also.[52]. Increased fibrosis post neoadjuvant chemotherapy has been suggested as one of the factors responsible for decreasing postoperative fistula rates in the neoadjuvant protocol.[53,54].In addition, it is suggested that delayed surgery in the neoadjuvant group can increase inflammatory changes due to prolong pancreatic ductal obstruction. [49].

Our aim was to analyze the effect of neoadjuvant chemoradiation on subsequent post-operative fistula rates by updated meta-analysis including all the recent publications in the last 10 years. We also analyzed other components of intraoperative fistula risk score like gland texture, blood loss, pancreatic duct size. We included studies with pancreatic ductal adenocarcinoma only, so histology was similar in the neoadjuvant first and surgery first group.

Pancreatic fistula rates were significantly lower in the neoadjuvant group. [Risk Ratio 0.66 (95 % C.I. 0.55-0.81). However, there were significantly fewer numbers of the soft pancreas and more mean blood loss in the neoadjuvant group. It shows that inflammatory changes post neoadjuvant therapy may help in decreasing intraoperative fistula risk score and may be responsible for decrease post-operative fistula risk score. Only two studies mentioned mean pancreatic diameter in their analysis, there was no significant difference in pancreatic duct diameter in both groups. PREOPANC trial [49] failed to show any difference in inflammatory changes between neoadjuvant and surgery first groups, but as discussed above our meta-analysis clearly showed that inflammatory changes were more prominent in the neoadjuvant group.

We also analyzed other confounding factors like the number of borderline resectable tumors and complication rates. Which showed no significant difference between two groups, further confirming the impact of neoadjuvant chemoradiotherapy on subsequent pancreatic fistula.

Our meta-analysis included all the recent publications regarding the impact of neoadjuvant treatment on subsequent postoperative pancreatic fistula and to our knowledge, this is the first meta-analysis that also included all the components of the fistula risk score. As, it is difficult to design randomized control trials in oncology keeping parameters like post-operative pancreatic fistula as the primary outcome, so such meta-analysis helps in obtaining more reliable shreds of evidence about the impact of neoadjuvant treatment on post-operative pancreatic fistula.

The majority of patients in the meta-analysis were having borderline resectable pancreatic cancer. Some centers are researching the impact of neoadjuvant chemotherapy on resectable pancreatic cancers.[55] The beneficial role of neoadjuvant treatment on subsequent clinically significant pancreatic fistula will make the case of using neoadjuvant chemoradiotherapy in this subgroup stronger.

There are certain limitations of this review heterogeneity was moderate to high in our analysis, though we used a random-effect model. There was also a single randomized control trial was available to include in this meta-analysis. As shown in figure 2, the risk of bias was high in some studies. As shown via funnel plots in supplement figure 1, we cannot entirely rule out the risk of publication bias also.

In, conclusion neoadjuvant treatments may be responsible for a lower rate of clinically significant pancreatic fistula after subsequent surgery, most likely due to its inflammatory response.

## Data Availability

data will be provided on demand.

## High quality

Further research is very unlikely to change our confidence in the estimate of effect.

## Moderate quality

Further research is likely to have an important impact on our confidence in the estimate of effect and may change the estimate.

## Low quality

Further research is very likely to have an important impact on our confidence in the estimate of effect and is likely to change the estimate.

## Very low quality

We are very uncertain about the estimate.

