## Supplementary material for "Impact of neoadjuvant chemoradiotherapy on post-operative clinically significant pancreatic fistula – A systemic review and updated meta-analysis": Supplmental Figure 1

(a)
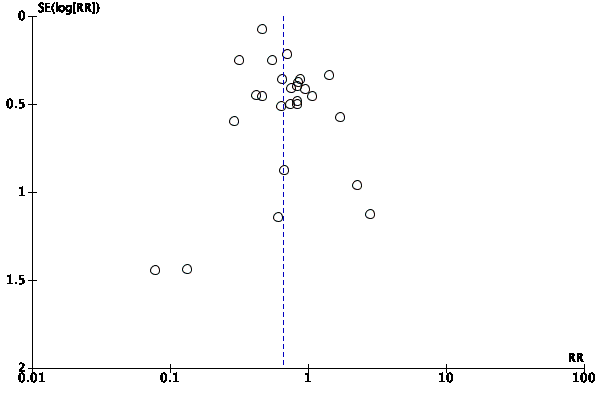

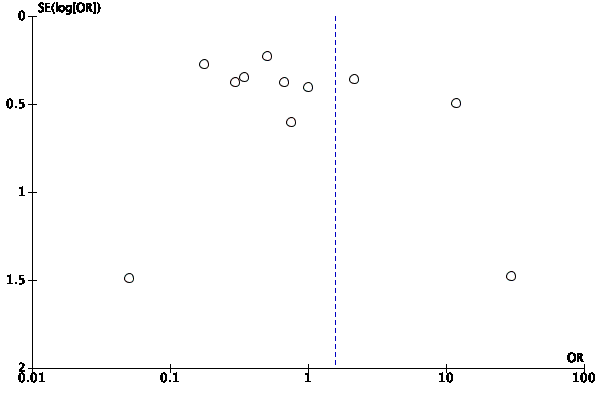
(b)

(c)
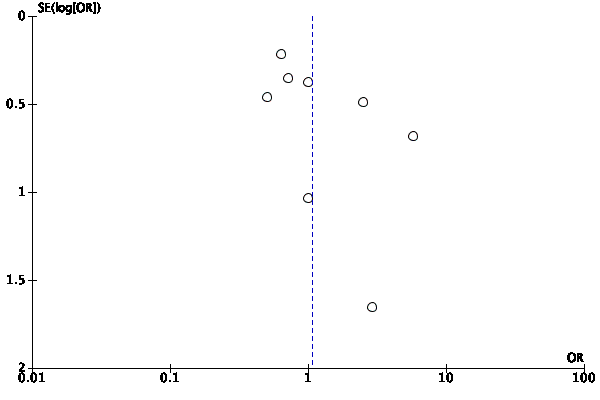

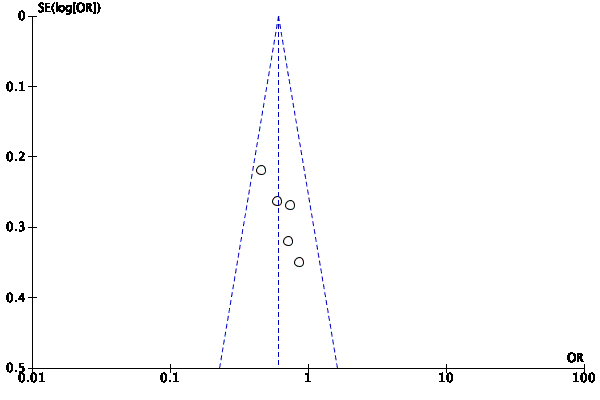

(d)

(e)
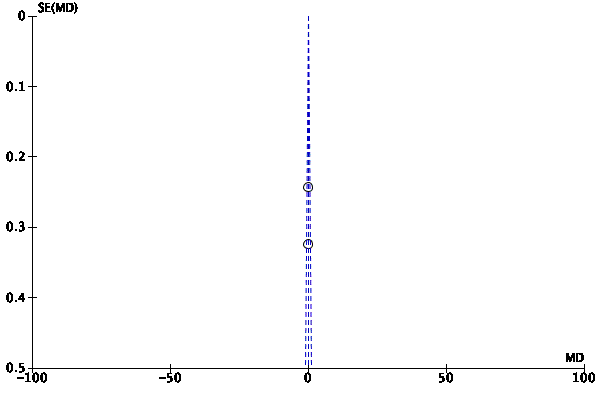

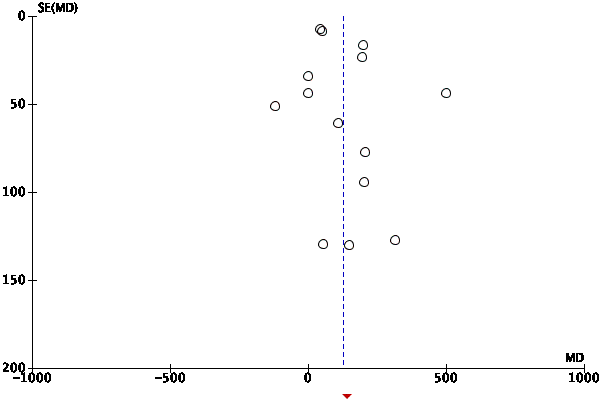

(f)

(g)
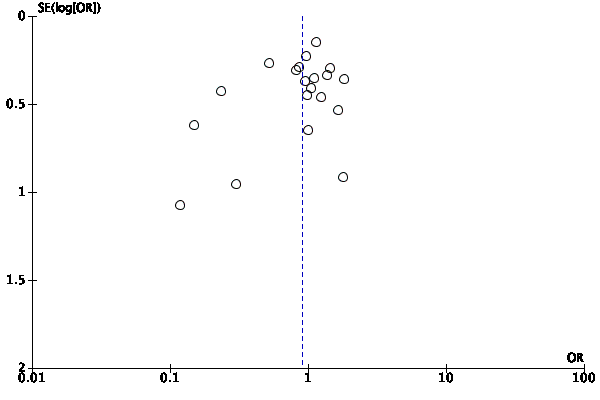

Supplement Figure1 : Funnel plot of comparison: (a) Neoadjuvant chemoradiotherapy vs direct surgery in pancreatic cancer for postoperative pancreatic fistula. (b) borderline resectable. (c) Distal pancreatectomy (d) Soft Pancreas (e) Pancreatic duct diameter (f) Blood loss (g) Complications.
